# To what extent could eliminating racial discrimination reduce inequities in mental health and sleep problems among Aboriginal and Torres Strait Islander children?

**DOI:** 10.1101/2023.11.23.23298938

**Authors:** Naomi Priest, Shuaijun Guo, Rushani Wijesuriya, Catherine Chamberlain (Palawa), Rosemary Smith (Ngarabul), Sharon Davis (Bardi Kija), Katherine Thurber, Janine Mohamed (Narrunga Kaurna), Margarita Moreno-Betancur

## Abstract

**Background:** Racism is a fundamental cause of health inequities for Aboriginal and Torres Strait Islander children. We aimed to examine the potential to reduce inequities in Aboriginal and Torres Strait Islander children’s mental health and sleep problems through eliminating interpersonal racial discrimination.

**Methods:** We drew on cross-sectional data from the Speak Out Against Racism (SOAR; N=2818) and longitudinal data from the Longitudinal Study of Australian Children (LSAC; N=8627). The SOAR survey was completed in 2017 and the LSAC followed children from 2004 to 2014 in the kindergarten cohort and from 2008 to 2018 in the birth cohort. Exposure was measured by Aboriginal and Torres Strait Islander status (Aboriginal and Torres Strait Islander or Anglo-European), as a proxy measure of structural racism (SOAR: 10-15 years; LSAC: 4-5 years). Mediator was measured by interpersonal racial discrimination (yes/no) (SOAR: 10-15 years; LSAC: 12-13 years). Outcomes were measured by mental health problems (yes/no) and sleep problems (yes/no) (SOAR: 10-15 years; LSAC: 14-15 years). An interventional effects approach was conducted, adjusting for baseline and intermediate confounders.

**Findings:** Aboriginal and Torres Strait Islander children had higher prevalence of mental health problems (SOAR: 40.1% versus 13.5%; LSAC: 25.3% versus 7.6%) and sleep problems (SOAR: 28.5% versus 18.4%; LSAC: 14.0% versus 9.9%) than their Anglo-European peers. Hypothetical interventions to eliminate Aboriginal and Torres Strait Islander children’s experiences of interpersonal racial discrimination could reduce up to 42.4% of mental health inequities (equivalent to 11.2% absolute reduction) and up to 48.5% of sleep inequities (equivalent to 4.7% absolute reduction).

**Interpretation:** Targeted policy interventions that eliminate racial discrimination against Aboriginal and Torres Strait Islander children have high potential to reduce inequities in mental health and sleep problems. Addressing racism and racial discrimination needs a multi-component and multi-level approach directed by Aboriginal and Torres Strait Islander communities.

**Funding:** National Health and Medical Research Council of Australia

**RESEARCH IN CONTEXT:** *Evidence before this study:* We searched four databases (Medline, PsycINFO, PubMed, and ERIC) via The University of Melbourne Library on 05, October 2023, for all publications since inception that used the interventional effects approach to quantify the extent to which intervening on racial discrimination could reduce disparities in health and wellbeing outcomes between Indigenous and non-Indigenous populations worldwide. This search did not identify any published studies, so we broadened our search to include studies with any population using a refined list of search terms: (“racism”) AND ((“interventional effects”) OR (“causal mediation”)) AND ((“health”) OR (“wellbeing”)). This search yielded a total of six studies, with all studies using causal mediation analysis to investigate racial and ethnic disparities (e.g., Black-White, Asian-White, Hispanic-White) in a range of health outcomes including mortality, preterm birth, substance misuse, and dementia among US adults. Mediators examined included hospital type, maternal cardiometabolic risk factors, area deprivation index, psychological distress, racial discrimination in medical settings, and systemic inflammation, with the proportion mediated ranging from 1.5% to 65.8%. However, no studies were identified that investigated the role of interpersonal racial discrimination in mediating racial and ethnic disparities in health among children.

*Added value of this study:* This study is the first to use an interventional effects approach to estimate the extent to which intervening on interpersonal racial discrimination could reduce inequities in mental health and sleep problems among Aboriginal and Torres Strait Islander children. We used both cross-sectional and longitudinal surveys to examine our research question, allowing us to explore common developmental associations within different samples across different study designs. We found that eliminating everyday experiences of racial discrimination could reduce almost half of the inequities in mental health (up to 42.4%) and sleep difficulties (up to 48.5%) experienced by Aboriginal and Torres Strait Islander children compared to their Anglo-European peers.

*Implications of all the available evidence:* Our findings confirm the well-established relationship between exposure to racism and poor mental health and sleep problems among Aboriginal and Torres Strait Islander children. The consistent benefit observed across cross-sectional and longitudinal studies suggests that targeted policy interventions that eliminate interpersonal racial discrimination have high potential to reduce inequities in mental health and sleep problems experienced by Aboriginal and Torres Strait Islander children. Critically, interpersonal racial discrimination is only one expression of the wider system of racism that impacts Aboriginal and Torres Strait Islander children. Multi-component and multi-level anti-racism actions directed by Aboriginal and Torres Strait Islander communities are needed to address racism in all its forms to achieve health equity and to realise fundamental human rights.

## INTRODUCTION

In Australia, for generations Aboriginal and Torres Strait Islander peoples have lived on, and from the land, nurturing the health and wellbeing of Aboriginal and Torres Strait Islander children through strong cultures, languages and systems of kinship and community care.^1^ Evidence shows that Aboriginal and Torres Strait Islander children were likely physically, socially and emotionally healthier than most European children at the time of colonization in 1788.^2^ Racism arrived in Australia at colonisation. Settler-colonists racialised and dehumanised Aboriginal and Torres Strait Islander peoples, considering them a culturally inferior and primitive race to legitimize and excuse systemic violence, genocide, forced removal from lands and suppression of cultural practices.^3^ Current differences observed across a range of social and health indicators between Aboriginal and Torres Strait Islander peoples and non-Indigenous people are not due to biology or race; they are the historical and ongoing effects of settler-colonialism and racism.^2^

Racism is an ideology and system of oppression that categorises and stratifies social groups into ‘races’ and then devalues and disadvantages those groups considered inferior and advantages those considered superior, differentially allocating to them valued societal power, resources and opportunities.^4^ Structural racism is the connected, reinforcing system of racialised rules, laws, policies, and regulations that are embedded in institutions such as healthcare, education, housing, justice, banking, and the media.^5^ Structural racism impacts multiple factors that lead to socioeconomic inequities, including access to education and employment, financial security, housing, and neighbourhood quality and safety. Directly and indirectly, including through socioeconomic pathways, structural racism influences exposure to physical, chemical and psychosocial stressors that affect risks for poor physical and mental health, social and emotional wellbeing, and health behaviors such as tobacco and alcohol use, dietary intake and physical activity.^6^ Cultural and ideological racism reinforce structural racism, with societal values and beliefs privileging and protecting Whiteness and White social and economic power.^7^ This includes framing populations as inferior or vulnerable in order to justify policies and systems that reinforce and embed inequities, for example, historical and contemporary policies of forced removal of Aboriginal and Torres Strait Islander children from their families (the Stolen Generations).^8^

Structural racism also manifests as interpersonal racial discrimination, an everyday form of racism expressed between individuals. Interpersonal racial discrimination enacts structural racism and reinforces racist ideologies - and is itself a powerful stressor and contributor to poor health and health inequities through mental health, behavioral and biological pathways.^9^ Aboriginal and Torres Strait Islander peoples have long said that interpersonal racial discrimination has harmful health consequences. Findings from systematic reviews and meta-analyses support this claim, with strong empirical evidence of negative health effects throughout life.^10,11^

Children are particularly sensitive to the negative effects of interpersonal racial discrimination with childhood developmentally, biologically and socially significant.^12^ Childhood sets the foundation for lifelong health and wellbeing with many chronic conditions including mental illness^13^ and cardiovascular disease^14^ shown to have early life origins. Childhood experiences of interpersonal racial discrimination are associated with negative effects on health in childhood and adulthood.^10,15^ Extant evidence among Aboriginal and Torres Strait Islander children aligns with international findings, documenting associations between childhood interpersonal racial discrimination and poor health, including mental health and sleep difficulties.^16^ However, the extent to which intervening on racial discrimination would reduce inequities in health experienced by Aboriginal and Torres Strait Islander children compared to their non-Indigenous peers remains unknown. To our knowledge no study internationally has estimated the extent to which intervening on racial discrimination would reduce inequities in Indigenous child health. Eliminating racism is a longstanding priority for Aboriginal and Torres Strait Islander peoples. Although addressing racism as a critical public health threat is increasingly recognised internationally,^17^ including for children,^18^ progress remains slow.

In this study, we aimed to quantify the potential benefit of intervening to eliminate interpersonal racial discrimination experiences among Aboriginal and Torres Strait Islander children to reduce inequities in mental health and sleep problems. We applied a new and novel statistical approach – causal mediation analysis based on the interventional effects approach (i.e., the evaluation of ‘what if’ intervention scenarios).^19^ We focused on mental health as the leading disease group contributing to disease burden in the Aboriginal and Torres Strait Islander population. The study was collaboratively developed between Aboriginal and non-Indigenous researchers from formation of research questions through to interpretation and discussion of findings.

## METHODS

### Data sources

We drew on both cross-sectional and longitudinal data from Australian large-scale population surveys to address the research question. Our purpose in bringing together these two studies is to examine our research question in two independent samples, allowing us to explore common developmental associations across different study designs.^20^

#### Speak Out Against Racism (SOAR)

SOAR is a large scale population-representative cross-sectional study on racism and mental health that was completed by 4664 primary and secondary students aged 10-15 years across 23 schools in New South Wales and Victoria in 2017.^21^ In brief, a stratification sampling method balanced on the available school characteristics was used to ensure the sample was representative of the overall school population in NSW and Victoria, with an oversampling of schools with higher proportions of Aboriginal and Torres Strait Islander students. Ethics approval of Speak Out Against Racism (SOAR) was obtained from the Australian National University Human Research Ethics Committee. Approval for researchers to approach schools to request participation in research was then sought from the Victorian and New South Wales Education Departments. Permission was obtained from each participating school principal, with parent opt-out consent and student assent.

#### Growing Up in Australia: The Longitudinal Study of Australian Children (LSAC)

LSAC is a nationally representative study comprised of two cohorts of Australian children: a birth cohort (B-cohort) of 5107 infants; and a kindergarten cohort (K-cohort) of 4983 four-year-olds. The study commenced in May 2004. In short, a two-stage clustered design was employed, where first the postcodes (i.e. the clusters) were randomly sampled, followed by randomly sampling children within each cluster, to select a sample that was broadly representative of the Australian child population except those living in remote areas (as some remote postcodes were excluded in stage one due to high cost of data collection).^22^ To ensure adequate statistical power to estimate meaningful associations for Aboriginal and Torres Strait Islander children, we drew on aligned data from both the B-cohort and K-cohort (see Supplementary file 1). LSAC was approved by the Australian Institute of Family Studies Human Research Ethics Review Board (ID 13-04).

### Measures

Our conceptual model shown in Figure 1 visually represents the hypothesized causal processes from structural to children’s mental health and sleep problems, informed by current knowledge (see Supplementary file 2). This model depicts the pathways from Aboriginal and/or Torres Strait Islander status – as a proxy for structural racism and associated exposures not as an indicator of innate, biological vulnerability^23^– to mental health and sleep problems, via interpersonal racial discrimination as the intervention of interest. Figure 1 was used to guide the selection of variables (Table 1) and inform the analytic approach.

**Figure 1.**
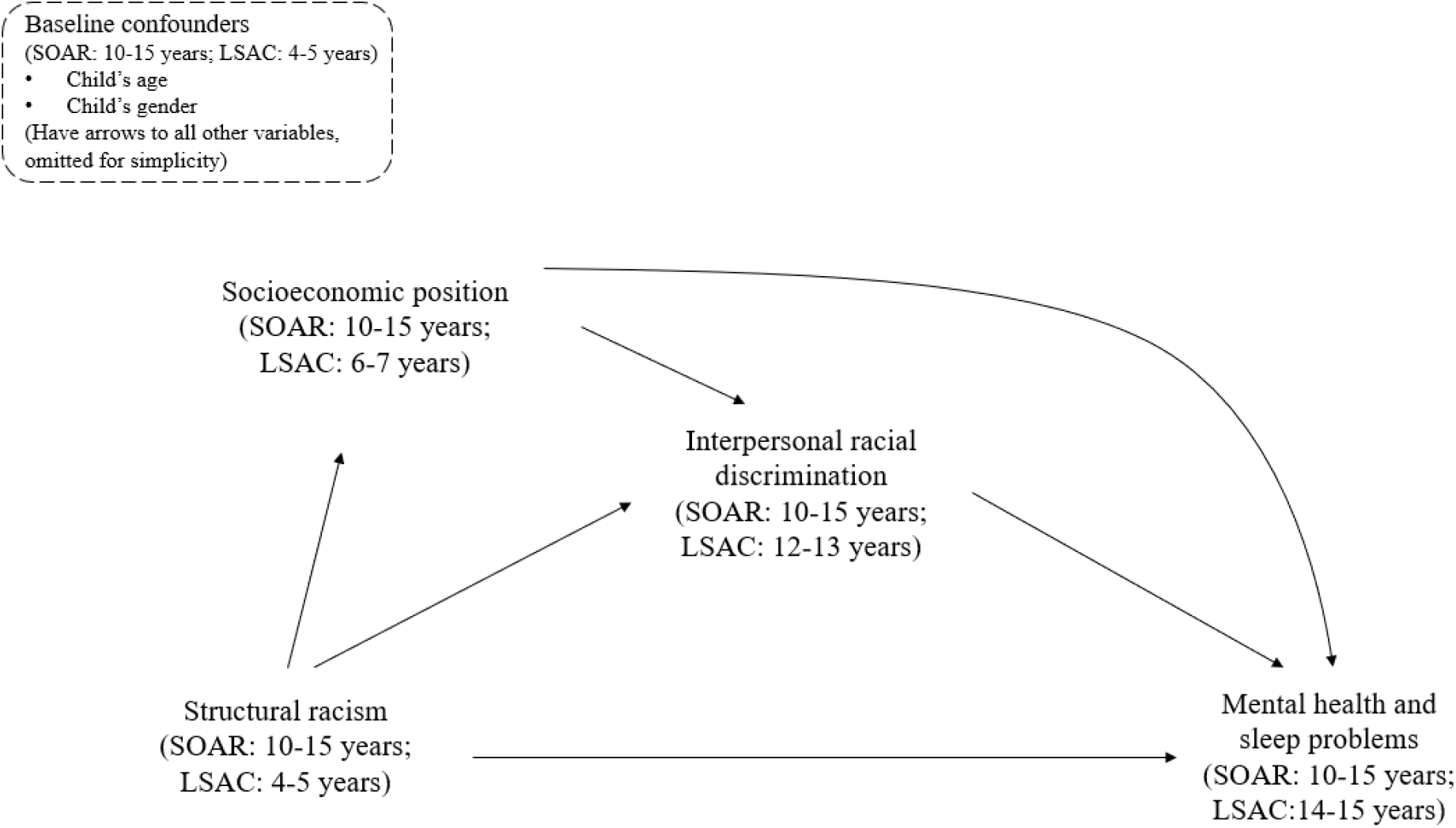
Conceptual model that depicts the relationship between Aboriginal and Torres Strait Islander status (as proxy for structural racism exposure rather than representing race and ethnicity as biological or innate constructs) and mental health and sleep problems via interpersonal racial discrimination.

**Table 1.** Measurement of exposure, mediator, outcomes and potential confounders.

| Variable | Measurement details |  |
| --- | --- | --- |
|  | SOAR | LSAC |
| **Exposure** |  |  |
| *Structural racism* | Aboriginal and Torres Strait Islander status and ethnicity in SOAR were captured at 10-15 years using a self-reported measure with categories developed for the study, given ethnicity beyond Aboriginal and Torres Strait Islander status is not routinely collected in Australia.<sup>21</sup> In this study, the SOAR sample was restricted to those from Aboriginal and Torres Strait Islander background (n=409) and Anglo-European backgrounds (n=2409). | Aboriginal and Torres Strait Islander and ethnicity in LSAC were derived from three indicators reported by the child's primary carer at 4-5 years: country of birth (both child and parents), language spoken at home (both child and parents), and Aboriginal and Torres Strait Islander status (child). We created three mutually exclusive categories: Aboriginal and/or Torres Strait Islander; ethnic minority (non-White and not Aboriginal and/or Torres Strait Islander) and Anglo-European following previous approaches.<sup>30</sup> Similarly, we restricted the LSAC sample to those from Aboriginal and Torres Strait Islander (n=417) and Anglo-European backgrounds (n=8210). |
| **Mediator** |  |  |
| *Interpersonal racial discrimination* | Direct racial discrimination in SOAR was measured at 10-15 years using 10 items drawn from the Adolescent Discrimination Distress Index (ADDI) together with two items used previously with diverse Australian school students.<sup>16</sup> Students responded to each item as 'this did not happen to me', 'once or twice', 'every few weeks', 'about once a week', 'several times a week or more'. A binary response ('This did not happen to me' versus 'once or twice/every few weeks/about once a week/several times a week or more') was created for each item. Students who answered 'yes' to any of the twelve items were considered as 'having ever experienced racial discrimination'. | Direct racial discrimination in LSAC was measured using three questions reported by the study child at 12-13 years. Children were asked whether in the last six months they had been treated unfairly or badly ('yes' or 'no') because of their language or accent, skin color, or cultural backgrounds. Children who answered 'yes' to any of the three questions were considered as 'having ever experienced racial discrimination'. |
| **Outcomes** |  |  |
| *Mental health problems* | Children's mental health problems were assessed using the Strengths and Difficulties Questionnaire (SDQ), which is a brief validated | The SDQ was also used in LSAC to measure children's mental health problems. Child-report SDQ ratings at 14-15 years were used in LSAC. Aboriginal and |
|  | SOAR | LSAC |
|  | screening measure of behavioral and emotional problems for 3-16 year olds. The SDQ total score of socioemotional difficulties is a sum of scores on 20 items, with higher scores representing poorer mental health. Student-report SDQ ratings at 10-15 years were used in SOAR. Given that the SDQ is not conceptually equivalent between Aboriginal and Torres Strait Islander children and non-Indigenous children,<sup>31</sup> Aboriginal and Torres Strait Islander children were coded as having elevated mental health symptoms if they scored 16-40 and non-Indigenous children if they scored 20-40. | Torres Strait Islander children were coded as having elevated mental health symptoms if they scored 16-40 and non-Indigenous children if they scored 20-40. |
| *Sleep problems* | Sleep problems at 10-15 years in SOAR were measured using the item “During the past four weeks, how often did you awake during your sleep time and have trouble falling back to sleep again?”, coded as “no” (none/a little, some/a good bit of the time) versus “yes” (most /all of the time). | Sleep problems at 14-15 years in LSAC were measured using the item “During the last month, how well do you feel you have slept in general?”, with sleep problems coded as “no” (very well/fairly well) versus “yes” (fairly badly/very badly). |
| **Confounders** |  |  |
| *Baseline confounders* | Two baseline confounders at 10-15 years were measured in SOAR: child’s age (continuous) and child’s sex (male/female). | Two baseline confounders at 4-5 years were measured in LSAC: child’s age (continuous) and child’s sex (male/female). |
| *Intermediate confounder* | Socioeconomic position was treated as an intermediate confounder, given it is affected by structural racism. Socioeconomic position at 10-15 years in SOAR was measured at the school level using the Index of Community Socio-Educational Advantage. In keeping with previous studies,<sup>32</sup> we used the bottom 25<sup>th</sup> percentile to indicate low socioeconomic position. | Socioeconomic position at 6-7 years in LSAC was measured at the family level using a composite of each parent’s self-reported annual income, highest education and occupation level. Following previous approaches,<sup>32</sup> socioeconomic position was dichotomized using the bottom 25<sup>th</sup> percentile to indicate low socioeconomic position. We also conducted a sensitivity analysis in LSAC, where there was broader data available, including five additional intermediate confounders at 6-7 years (household member mental health, household member substance use, family violence, child disability and remoteness) (see Supplementary file 3 for details). |

### Statistical analysis

Descriptive statistics were calculated for all variables of interest using counts and proportions for binary variables and using means and standard deviations for continuous variables in the total sample and stratified by Aboriginal and Torres Strait Islander status. Preliminary analyses were then conducted to examine the pathways depicted in Figure 1. We used generalised estimating equations (GEE) to account for the cluster correlations due to schools (SOAR) and postcodes (LSAC) in the data. Specifically, GEE with log-Poisson link and an exchangeable correlation matrix were used to estimate risk ratios representing the unadjusted and adjusted (conditional) associations between Aboriginal and Torres Strait Islander status and each outcome (mental health and sleep problems), between Aboriginal and Torres Strait Islander status and racial discrimination, and between racial discrimination and each outcome. Descriptive statistics and preliminary analyses were conducted using Stata 17.0.

#### Causal mediation analysis

For each outcome (mental health and sleep problems), we conducted a causal mediation analysis using an extended g-computation approach outlined below to estimate (1) the overall inequities in the prevalence of each outcome (mental health and sleep problems) between Aboriginal and Torres Strait Islander children and those from Anglo-European background, and (2) the benefit (in terms of reducing outcome prevalence) of a hypothetical intervention eliminating racial discrimination (i.e., a maximum benefit intervention scenario) in Aboriginal and Torres Strait Islander children. For this purpose, we followed the framework outlined by Moreno-Betancur et al.^19^, defining interventional mediation effects that map to the target trial, i.e. the randomized trial that would have been ideally conducted to assess the impact of such a hypothetical intervention. Analyses using interventional effects are increasingly used in health inequities research and are ideal for contexts where data on actual, well-defined interventions already implemented in the community are not available.^24^

Effects were estimated using an extended g-computation estimation procedure, which uses a series of richly specified (with interactions) regression models for any intermediate confounders, the mediator and the outcome. In both primary and sensitivity analyses, the overall inequities (with no mediator intervention) were estimated by comparing the mean outcome under exposure and no exposure, estimated using the outcome models detailed in Supplementary file 3. The reductions in prevalence of each outcome that would be achieved by an intervention that would completely eliminate racial discrimination experienced by Aboriginal and Torres Strait Islander children were estimated by setting the values of the mediator under exposure to zero. From this, we could estimate the inequities in each outcome that would remain after intervening to eliminate racial discrimination in Aboriginal and Torres Strait Islander children. Standard errors for all estimates were computed using a clustered bootstrap procedure. All causal mediation analyses were implemented using R Statistical Software 4.2.2.

#### Missing data

The proportion of children with missing data across all study variables was 12.1% and 41.4% in SOAR and LSAC, respectively. Multiple imputation was used to reduce bias due to incomplete records for preliminary and causal mediation analyses. Multiple imputation was implemented using multivariate imputation by chained equations separately in each study. We imputed continuous and binary variables using univariate linear and logistic regression models respectively using Stata 17.0. The imputation models included all study variables as well as all two-way interactions amongst exposure, mediator, outcome, baseline and intermediate confounders as indicated by the regression models used in extended g-computation for the causal mediation analysis (see Supplementary file 4). Clustering by school in SOAR and by residential postcodes in LSAC was accounted for in the imputation procedure by including a categorical cluster membership variable as a predictor in the univariate imputation models. Based on the percentage of missing data, we imputed twenty datasets for SOAR and forty for LSAC. Post-imputation, the analyses were conducted in each of the imputed dataset and the results were combined using Rubin’s rules to obtain the final imputed estimates of the parameters of interest.

## RESULTS

### Sample characteristics

Participant characteristics are summarized in Table 2. SOAR had a higher proportion of Aboriginal and Torres Strait Islander children than LSAC (14.5% vs 4.8%). Around half (50.1%) of Aboriginal and Torres Strait Islander children in SOAR experienced racial discrimination and 26.0% in LSAC. At outcome assessment, Aboriginal and Torres Strait Islander children in SOAR had higher prevalence of elevated mental health symptoms (40.1% versus 25.3%) and sleep problems (28.5% versus 14.0%) than those in LSAC.

**Table 2.**
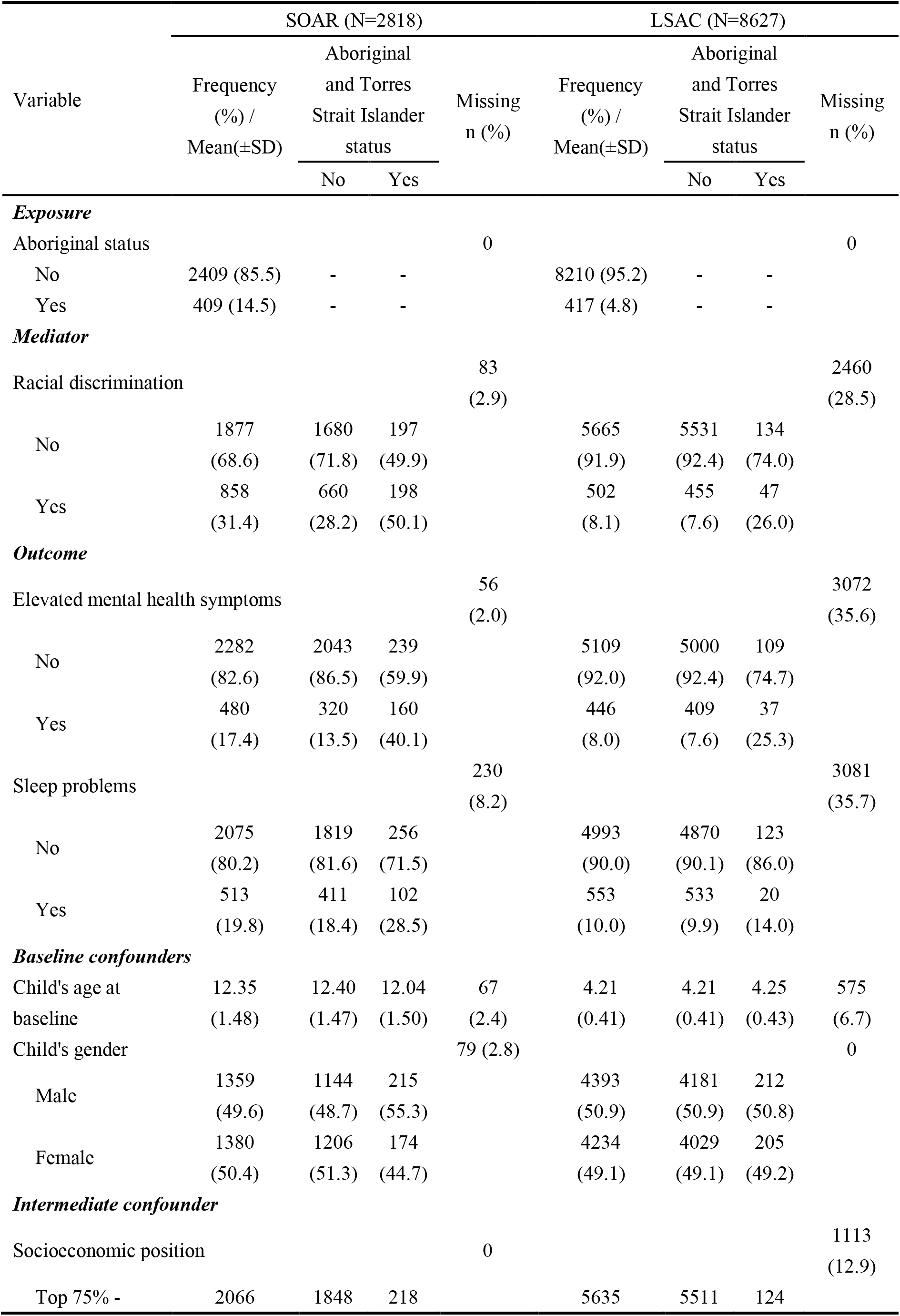
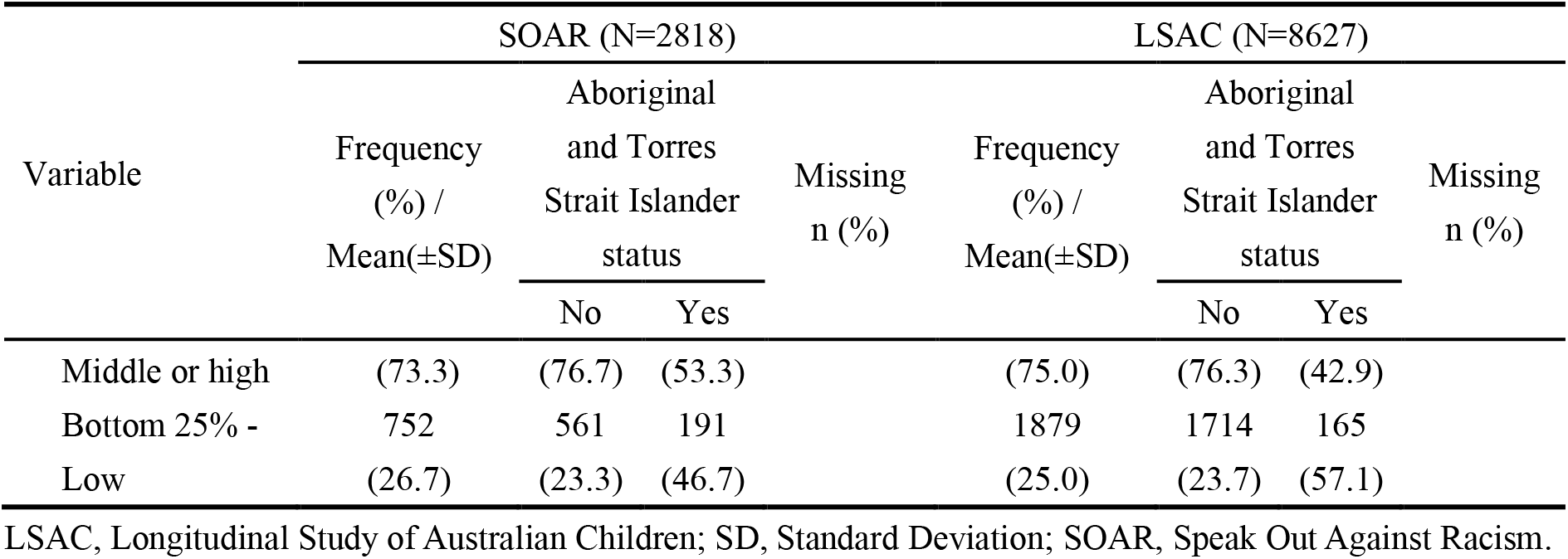
Descriptive information for all study variables in SOAR and LSAC samples. Observed data are shown.

### Associations between Aboriginal and Torres Strait Islander status, racial discrimination, and mental health and sleep problems

Table 3 shows the preliminary results regarding associations between the main variables of interest. Aboriginal and Torres Strait Islander children had higher risk of experiencing racial discrimination (SOAR: RR=1.69, 95% CI=1.47 to 1.95; LSAC: RR=3.35, 95% CI=2.54 to 4.42), elevated mental health symptoms (SOAR: RR=2.89, 95% CI=2.27 to 3.69; LSAC: RR=3.45, 95% CI=2.70 to 4.43) and sleep problems (SOAR: RR=1.42, 95% CI=1.09 to 1.85; LSAC: RR=1.56, 95% CI=1.05 to 2.32) than their peers from Anglo-European backgrounds, after adjusting for gender and age. Children who experienced racial discrimination had higher risk of elevated mental health symptoms (SOAR: RR=2.19, 95% CI=1.81 to 2.66; LSAC: RR=2.27, 95% CI=1.81 to 2.84) and sleep problems (SOAR: RR=1.58, 95% CI=1.35 to 1.84; LSAC: RR=1.59, 95% CI=1.22 to 2.08) than those who did not, after adjusting for gender, age, Aboriginal and Torres Strait Islander status and socioeconomic position. These findings provide support for the detrimental effect of racial discrimination on children’s mental health and sleep problems.

**Table 3.** Generalized estimating equations examining the associations between Aboriginal status, racial discrimination, and mental health and sleep problems, based on multiple imputation in each study (SOAR, N=2818 and LSAC, N=8627).

| Models | SOAR (RR and its 95% CI) |  | LSAC (RR and its 95% CI) |  |
| --- | --- | --- | --- | --- |
|  | Unadjusted | Adjusted* | Unadjusted | Adjusted* |
| ***Association with elevated mental health symptoms*** |  |  |  |  |
| Aboriginal and Torres Strait Islander status (Ref=no) | 2.87 (2.24, 3.68) | 2.89 (2.27, 3.69) | 3.46 (2.7, 4.44) | 3.45 (2.70, 4.43) |
| Racial discrimination (Ref=no) | 2.46 (2.02, 3.00) | 2.19 (1.81, 2.66) | 2.74 (2.2, 3.41) | 2.27 (1.81, 2.84) |
| ***Association with sleep problems*** |  |  |  |  |
| Aboriginal and Torres Strait Islander status (Ref=no) | 1.40 (1.08, 1.82) | 1.42 (1.09, 1.85) | 1.56 (1.05, 2.33) | 1.56 (1.05, 2.32) |
| Racial discrimination (Ref=no) | 1.60 (1.36, 1.88) | 1.58 (1.35, 1.84) | 1.64 (1.28, 2.10) | 1.59 (1.22, 2.08) |
| ***Association with racial discrimination*** |  |  |  |  |
| Aboriginal and Torres Strait Islander status (Ref=no) | 1.72 (1.49, 1.98) | 1.69 (1.47, 1.95) | 3.36 (2.55, 4.44) | 3.35 (2.54, 4.42) |

CI, confidence interval; LSAC, Longitudinal Study of Australian Children; Ref, reference group; RR, risk ratio; SOAR, Speak Out Against Racism. \* Adjusted confounders were child's age and gender. Models for the association between racial discrimination and each outcome were additionally adjusted for Aboriginal and Torres Strait Islander status and socioeconomic position.

### Extent to which a hypothetical intervention eliminating racial discrimination reduces inequities

Table 4 displays the results from the causal mediation analysis using the interventional effects approach. In terms of the overall inequities in each outcome between Aboriginal and Torres Strait Islander children and their Anglo-European peers, we found that the absolute difference in the prevalence of elevated mental health symptoms was 26.4% (95% CI: 21.4% to 31.3%) in SOAR, adjusted for age and gender, while for sleep problems it was 9.7% (95% CI: 3.3% to 16.1%). Similarly, in LSAC, the absolute difference was 20.1% (95% CI 13.4%, 26.9%) in the prevalence of elevated mental health symptoms and 6.2% (95% CI: 0.0% to 12.4%) in sleep problems for Aboriginal and Torres Strait Islander children compared with their Anglo-European peers, after adjusting for age and gender.

**Table 4.** Results from causal mediation analysis: Estimated effects on the prevalence of mental health and sleep problems by hypothetical interventions eliminating racial discrimination in Aboriginal and Torres Strait Islander children, using multiply imputed data for the full cohorts in SOAR and LSAC.

| Effect | SOAR (N=2818) |  |  | LSAC (N=8627) |  |  |
| --- | --- | --- | --- | --- | --- | --- |
|  | Estimate of absolute risk reduction (%)<br>95% CI | Proportion of inequities eliminated | Remaining inequities (%)<br>95% CI | Estimate of absolute risk reduction (%)<br>95% CI | Proportion of inequities eliminated | Remaining inequities (%)<br>95% CI |
| ***Mental health problems*** |  |  |  |  |  |  |
| Adjusted prevalence difference (overall inequities) | 26.4<br>(21.4, 31.3) | - | - | 20.1<br>(13.4, 26.9) | - | - |
| Risk reduction from intervening on racial discrimination | 11.2<br>(5.6, 16.8) | 42.4 | 15.2<br>(9.0, 21.3) | 5.3<br>(0, 10.6) | 26.4 | 14.8<br>(6.6, 23.0) |
| ***Sleep problems*** |  |  |  |  |  |  |
| Total adjusted marginal prevalence difference | 9.7<br>(3.3, 16.1) | - | - | 6.2<br>(0, 12.4) | - | - |
| Risk reduction from intervening on racial discrimination | 4.7<br>(0.3, 9.2) | 48.5 | 5.0<br>(0, 12.0) | -0.3<br>(-4.4, 3.7) | 0 | 6.5<br>(-0.8, 13.9) |

CI, confidence interval. Baseline confounders controlled for were child's age and gender. Intermediate confounder controlled for was socioeconomic position.

In SOAR, we estimated that a hypothetical intervention eliminating racial discrimination experienced by Aboriginal and Torres Strait Islander children would result in a 11.2% (95%CI : 5.6% to 16.8%) and 4.7% (95% CI: 0.3% to 9.2%) absolute reduction in prevalence of elevated mental health symptoms and sleep problems in Aboriginal and Torres Strait Islander children. These estimates correspond to 42.4% and 48.5% inequities eliminated in mental health and sleep problems respectively. In LSAC, we found that a hypothetical intervention eliminating racial discrimination experienced by Aboriginal and Torres Strait Islander children would result in an absolute reduction of 5.3% (95%CI: 0% to 10.6%) in the prevalence of elevated mental health symptoms, corresponding to 26.4% mental health inequities eliminated. The reduction in the prevalence of sleep problems was negligible (-0.3%, 95% CI -4.4% to 3.7%). Estimates in LSAC remained largely unchanged when the causal mediation analysis approach also modelled the five additional intermediate confounders (Supplementary file 2). In SOAR, almost half of the existing inequities were reduced after the hypothetical intervention on racial discrimination. However, in LSAC, large inequities remain after the hypothetical intervention on racial discrimination.

## DISCUSSION

### Summary of key findings

Using both cross-sectional and longitudinal study designs, we estimated the potential benefit of hypothetical interventions that would eliminate interpersonal racial discrimination to reduce inequities in mental health and sleep problems among Aboriginal and Torres Strait Islander children. Our findings confirmed the large existing inequities in mental health and sleep problems between Aboriginal and Torres Strait Islander children and their Anglo-European peers.^16,25^ Findings were consistent in SOAR and LSAC regarding the potential benefit of eliminating racial discrimination to reduce mental health inequities among Aboriginal and Torres Strait Islander children. We also observed substantial benefits in reducing inequities in sleep problems in SOAR.

Consistent with previous findings,^26^ we found that interpersonal racial discrimination interventions had positive effects on children’s mental health and sleep outcomes. Using data from the Longitudinal Study of Indigenous Children, Shepherd et al.^27^ used the population attributable risk (PAR) as a measure of how much disease could be averted if interpersonal racial discrimination was eliminated. They found that racial discrimination accounted for 16.2% and 19.1% of the PAR for mental health problems and sleep difficulties respectively. To our knowledge, there have been no studies globally that specifically quantify the extent to which intervening on interpersonal racial discrimination could reduce inequities in mental health and sleep problems among Indigenous children. Our findings add to current evidence regarding the substantial impact of racial discrimination on child health and child health inequities and reinforce calls to address racism and racial discrimination as a critical and urgent public health priority. We found that eliminating interpersonal racial discrimination could reduce close to half of the inequities in mental health and sleep difficulties experienced by Aboriginal and Torres Strait Islander children. To our knowledge no other factor has been found to explain such a substantial proportion of health inequities experienced by Aboriginal and Torres Strait Islander children. Using population attributable fractions (PAFs) Thurber et al^8^ recently showed that everyday racial discrimination could explain 47.4% of the overall gap in psychological distress between Indigenous and non-Indigenous adults. This further reinforces the substantial impact of racial discrimination on mental health inequities experienced by Aboriginal and Torres Strait Islander peoples. Urgent re-orientation of health policy, practice and research to address racism among Aboriginal and Torres Strait Islander children, their families and communities is required.

While the results suggest the potential benefits of eliminating racial discrimination, more than half of inequities in mental health and sleep problems remain between Aboriginal and Torres Strait Islander children and their Anglo-European peers after the hypothetical intervention. Structural racism can impact health through a range of direct and indirect pathways, with interpersonal racial discrimination only one of these.^6,7^ Notwithstanding the critical importance of addressing interpersonal racial discrimination, further action is needed across multiple strategies to address structural racism and to fully achieve health equity and human rights. We suggest that if racism in all forms was eliminated from society, there would be no remaining health inequities experienced by Aboriginal and Torres Strait Islander children at all.

We also found that the potential benefit of eliminating experiences of Aboriginal and Torres Strait Islander children’s racial discrimination appeared to be more prominent in SOAR than that in LSAC. There are two key differences that may explain these differences between SOAR and LSAC. First, SOAR had a more representative sample of Aboriginal and Torres Strait Islander children than LSAC. Second, SOAR used a 12-item instrument to measure interpersonal racial discrimination that captured a broad range of experiences with five response categories for each item, whereas LSAC asked children to answer yes or no using three general items (“because of their language or accent, skin color, or cultural backgrounds”) regarding racial discrimination. Further studies are needed to replicate our findings.

### Strengths and limitations

A key strength of this study is the replication of analyses in two representative samples of Australian children from both longitudinal and cross-sectional perspectives, enhancing confidence of our findings. Second, we used emerging causal mediation methods that allow us to draw meaningful and policy-relevant insights from existing high-quality observational data. Nevertheless, there are several limitations. First, measurement errors likely exist for structural racism and interpersonal racial discrimination. We rely on Aboriginal and Torres Strait Islander status as a proxy measure of structural racism, in the absence of additional measures that capture the complexity of social and institutional aspects of structural racism. Interpersonal racial discrimination measures used in the present study only capture a limited range of experience, particularly in LSAC. Therefore, the potential benefits were likely underestimated. Second, the hypothetical intervention that would be capable of achieving the potential benefits is ill-defined. Third, while we accounted for a host of baseline and intermediate confounders, unmeasured factors may still confound the associations, which could result in bias in our estimates.

### Implications and future directions

In a context where the COVID-19 pandemic and the Voice referendum has resulted in rising racism towards Aboriginal and Torres Strait Islander children and magnified the inequities they, their families and communities experience,^28^ actions to address racism and its impacts are urgent and essential. Our findings suggest eliminating interpersonal racial discrimination would have substantial benefits for reducing inequities in mental health and sleep problems among Aboriginal and Torres Strait Islander children. These findings from observational data should be triangulated with further empirical evidence, including anti-racism intervention programs in the real world.

Interpersonal racial discrimination is only the tip of the iceberg of the system of racism that impacts on Aboriginal and Torres Strait Islander children, families and communities. Further attention should be given to upstream structural racism interventions within settings such as healthcare, housing, employment, and addressing outstanding issues such as constitutional recognition and meeting human rights obligations.^29^ In a world where there is no structural racism, there would be no gap at all between Aboriginal and Torres Strait Islander children and their non-Indigenous peers. Future efforts are needed to address racism driven by Aboriginal and Torres Strait Islander communities.

## CONCLUSIONS

We found that eliminating interpersonal racial discrimination could reduce close to half of the inequities in mental health and sleep difficulties experienced by Aboriginal and Torres Strait Islander children compared to their Anglo-European peers. Attention to interpersonal racial discrimination without addressing the more upstream structural racism may produce few gains. Continued policy and practice efforts are needed driven by Aboriginal and Torres Strait Islander communities to address racism and its negative impact on Aboriginal and Torres Strait Islander children.

## Data Availability

All data used in this manuscript are available on application to the Longitudinal Study of Australian Children (https://dataverse.ada.edu.au/dataverse/lsac) and from the corresponding author on request on the Speak Out Against Racism.

## Contributors

NP conceptualized the initial idea for the study. NP, SG, RW, and MM-B contributed to the data analysis plan. SG and RW did the data analysis. MM-B provided statistical advice. RW and MM-B cross-checked coding and verified the data. SG did the literature review. NP and SG produced the initial draft of the manuscript. All authors were involved in refining the scope of the study, interpreting results and drafting the manuscript. All authors approved the final version of the manuscript, take responsibility for its content, and were responsible for the decision to submit the manuscript.

## Acknowledgments

This paper uses unit record data from Growing Up in Australia, the Longitudinal Study of Australian Children (LSAC) and the Speak Out Against Racism (SOAR). LSAC is conducted by the Australian Government Department of Social Services (DSS). The findings and views reported in this paper, however, are those of the authors and should not be attributed to the Australian Government DSS or any of DSS’ contractors or partners. DOI: 10.26193/F2YRL5. The authors would like to thank all schools and students participating in SOAR. The authors would like to thank Tania King for her work during the early stages of SOAR and research staff (Rebecca Moorhead, Sharon Moorhead, Brandi Fox, Meiliasari Meiliasari and Emma Whatman (Victoria); and Oishee Alam, Alexia Derbas, Katie Blair, Rosalie Atie and Zarlasht Sarwari (New South Wales)) who were involved in data collection. The authors would like to acknowledge the support of the Social Research Centre with data collection of SOAR. KT and MM-B are supported by the National Health and Medical Research Council of Australia.

## REFERENCES

1. McMahon M. Lotjpa-nhanuk: Indigenous Australian child-rearing discourses. La Trobe University; 2017.

2. Thomson N. Australian aboriginal health and health-care. Social Science & Medicine. 1984/01/01/ 1984;18(11):939-948. 10.1016/0277-9536(84)90264-8

3. Borch M. Rethinking the origins of terra nullius. Australian Historical Studies. 2001;32(117):222–239.

4. Bonilla-Silva E. Rethinking Racism - toward a structural interpretation. American Sociological Review. 1997;62(3):465–480. 10.2307/2657316

5. Gee GC, Hicken MT. Structural Racism: The Rules and Relations of Inequity. Ethn Dis. 2021;31(Suppl 1):293–300. doi:10.18865/ed.31.S1.293

6. Braveman PA, Arkin E, Proctor D, Kauh T, Holm N. Systemic And Structural Racism: Definitions, Examples, Health Damages, And Approaches To Dismantling. Health Aff (Millwood). Feb 2022;41(2):171–178. doi:10.1377/hlthaff.2021.01394

7. Michaels EK, Lam-Hine T, Nguyen TT, Gee GC, Allen AM. The Water Surrounding the Iceberg: Cultural Racism and Health Inequities. The Milbank Quarterly. 2023;101(3):768–814.

8. Thurber KA, Brinckley M-M, Jones R, et al. Population-level contribution of interpersonal discrimination to psychological distress among Australian Aboriginal and Torres Strait Islander adults, and to Indigenous–non-Indigenous inequities: cross-sectional analysis of a community-controlled First Nations cohort study. The Lancet. 2022;400(10368):2084–2094. doi:10.1016/S0140-6736(22)01639-7

9. Nazroo JY, Bhui KS, Rhodes J. Where next for understanding race/ethnic inequalities in severe mental illness? Structural, interpersonal and institutional racism. Sociology of health & illness. 2020;42(2):262–276. doi:10.1111/1467-9566.13001

10. Paradies Y, Ben J, Denson N, et al. Racism as a Determinant of Health: A systematic review and meta-analysis. PLoS One. 2015;10(9):e0138511. doi:10.1371/journal.pone.0138511

11. Kairuz CA, Casanelia LM, Bennett-Brook K, Coombes J, Yadav UN. Impact of racism and discrimination on physical and mental health among Aboriginal and Torres Strait islander peoples living in Australia: a systematic scoping review. BMC Public Health. Jul 3 2021;21(1):1302. doi:10.1186/s12889-021-11363-x

12. Gee GC, Walsemann KM, Brondolo E. A life course perspective on how racism may be related to health inequities. American Journal of Public Health. 2012-5 2012;102(5):967-974. doi:10.2105/AJPH.2012.300666

13. Kessler RC, Chiu WT, Demler O, Merikangas KR, Walters EE. Prevalence, severity, and comorbidity of 12-month DSM-IV disorders in the National Comorbidity Survey Replication. Archives Of General Psychiatry. Jun 2005;62(6):617–627. doi:10.1001/archpsyc.62.6.617

14. Jacobs DR, Woo JG, Sinaiko AR, et al. Childhood Cardiovascular Risk Factors and Adult Cardiovascular Events. New England Journal of Medicine. 2022;386(20):1877–1888. doi:10.1056/NEJMoa2109191

15. Priest N, Paradies Y, Trenerry B, Truong M, Karlsen S, Kelly Y. A systematic review of studies examining the relationship between reported racism and health and wellbeing for children and young people. Social Science & Medicine. 2013;95:115–127. doi:10.1016/j.socscimed.2012.11.031

16. Priest N, Chong S, Truong M, et al. Racial discrimination and socioemotional and sleep problems in a cross-sectional survey of Australian school students. Archives of Disease in Childhood. Jul 28 2020;105(11):1079–1085. doi:10.1136/archdischild-2020-318875

17. Devakumar D, Selvarajah S, Shannon G, et al. Racism, the public health crisis we can no longer ignore. Lancet. 2020;395(10242):e112–e113. doi:10.1016/s0140-6736(20)31371-4

18. Trent M, Dooley DG, Douge J, Section On Adolescent H, Council On Community P, Committee On A. The Impact of Racism on Child and Adolescent Health. Pediatrics. Aug 2019;144(2)doi:10.1542/peds.2019-1765

19. Moreno-Betancur M, Moran P, Becker D, Patton GC, Carlin JB. Mediation effects that emulate a target randomised trial: Simulation-based evaluation of ill-defined interventions on multiple mediators. Statistical Methods in Medical Research. 2021;30(6):1395–412. doi:10.1177/0962280221998409

20. O’Connor M, Spry E, Patton G, et al. Better together: Advancing life course research through multi-cohort analytic approaches. Advances in Life Course Research. 2022/09/01/ 2022;53:100499. 10.1016/j.alcr.2022.100499

21. Priest N, Chong S, Truong M, et al. Findings from the 2017 Speak Out Against Racism (SOAR) student and staff surveys. CSRM working paper *no.* 3/2019. 2019. https://csrm.cass.anu.edu.au/sites/default/files/docs/2019/8/CSRM-WP-SOAR_PUBLISH_1.pdf

22. Soloff C, Lawrence D, Johnstone R. LSAC Technical paper No. 1. Sample design. Australian Institute of Family Studies; 2005.

23. Lett E, Asabor E, Beltrán S, Cannon AM, Arah OA. Conceptualizing, Contextualizing, and Operationalizing Race in Quantitative Health Sciences Research. Annals of family medicine. Mar-Apr 2022;20(2):157–163. doi:10.1370/afm.2792

24. Jackson JW. Meaningful Causal Decompositions in Health Equity Research: Definition, Identification, and Estimation Through a Weighting Framework. Epidemiology. 2021;Publish Ahead of Print

25. Blunden S, Fatima Y, Yiallourou S. Sleep health in Indigenous Australian children: a systematic review. Sleep medicine. 2021;80:305–314.

26. Priest N, Alam O, Truong M, et al. Promoting proactive bystander responses to racism and racial discrimination in primary schools: a mixed methods evaluation of the ‘Speak Out Against Racism’ program pilot. BMC Public Health. 2021/07/21 2021;21(1):1434. doi:10.1186/s12889-021-11469-2

27. Shepherd CCJ, Li J, Cooper MN, Hopkins KD, Farrant BM. The impact of racial discrimination on the health of Australian Indigenous children aged 5–10 years: analysis of national longitudinal data. journal article. International Journal for Equity in Health. July 03 2017;16(1):116. doi:10.1186/s12939-017-0612-0

28. Coyne-Beasley T, Hill SV, Miller E, Svetaz MV. Health Equity and the Impact of Racism on Adolescent Health. Pediatrics. 2023;151(Supplement 1)

29. Dudgeon P, Bray A, Walker R. Mitigating the impacts of racism on Indigenous wellbeing through human rights, legislative and health policy reform. Medical Journal of Australia. 2023;218(5):203–205. doi:10.5694/mja2.51862

30. Priest N, King T, Bécares L, Kavanagh A, M. Bullying victimization and racial discrimination among Australian children. Article. American Journal of Public Health. 2016;106(10):1882–1884. doi:10.2105/AJPH.2016.303328

31. Thurber K, Walker J, Dunbar T, et al. Measuring child mental health, psychological distress, and social and emotional wellbeing in the longitudinal study of indigenous children. 2019.

32. Goldfeld S, Moreno-Betancur M, Gray S, et al. Addressing child mental health inequities through parental mental health and preschool attendance. Pediatrics. 2023;151(5)

