## Supplementary files for "To what extent could eliminating racial discrimination reduce inequities in mental health and sleep problems among Aboriginal and Torres Strait Islander children?"

**Supplementary file 1: Flowchart of LSAC participants**

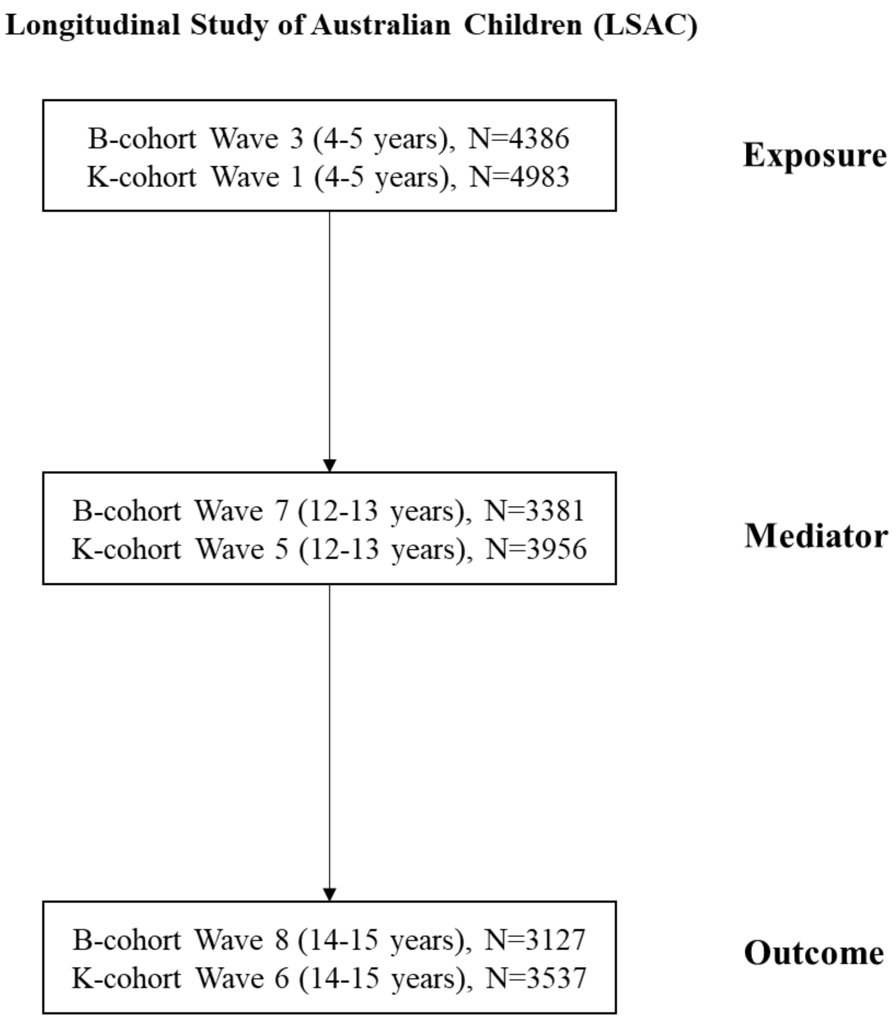


eFigure 1. Flowchart of LSAC participants

**Supplementary file 2.** Rationale for the pathways specified in the conceptual model

eTable 1. Justifications for the inclusion of variables and pathways specified in the conceptual model, including justification of variable cut-offs.

| Type of variable | Variable | Cut-off decision and citation | • Relationship with exposure | Relationship with mediator and intermediate confounder | Relationship with outcome |
|---|---|---|---|---|---|
| Exposure | Structural racism (Aboriginal and Torres Strait Islander status) | Yes versus no[1,2] | • - | • Aboriginal and Torres Strait Islander children are at high risk of experiencing racial discrimination compared with non-Indigenous children.[3]<br>• Aboriginal and Torres Strait Islander families have higher unemployment rates and earn lower household incomes than non-Aboriginal and Torres Strait Islander families.[4] (Due to settler colonisation and racism) | • Aboriginal and Torres Strait Islander children are more likely to have social and emotional difficulties than non-Indigenous children.[5,6]<br>• Compared with non-Indigenous children, Aboriginal and Torres Strait Islander children are more likely to experience sleep problems.[7] |
| Mediator | Interpersonal racial discrimination | Yes versus no[8] | • - | - | • Experiences of direct racial discrimination are associated with children's socioemotional and sleep problems.[9,10] |
| Outcome | Mental health problems | 0-15 versus 16-40 for Aboriginal and Torres Strait Islander children[11] and 0-19 versus 20-40 for non-Indigenous children[12] | • - | - | - |
| | Sleep problems | Yes versus no[13,14] | • - | - | - |
| Baseline confounder | Child's age | Continuous | • Age is a covariate that may influence the association between | Age is a covariate the may influence the association between socioeconomic position, racial | • Older children are more likely to experience mental health problems |

| | | | | | |
|---|---|---|---|---|---|
| | | | Aboriginal and Torres Strait Islander status and child mental health.[6] | discrimination and child mental health.[10] | than younger children.[15]<br>Age is associated with sleep problems in children.[16] |
| | Child's gender | Male versus female | • Gender is a covariate that may influence the association between Aboriginal status and child mental health.[6] | Gender is a covariate the may influence the association between socioeconomic position, racial discrimination and child mental health.[10] | • Boys are more likely than girls to experience mental health problems.[15]<br>Gender is associated with sleep problems in children.[16] |
| Intermediate confounder | Socioeconomic position | Bottom 25% versus top 75%[17] | • Socioeconomic position is a covariate that may influence the association between Aboriginal and Torres Strait Islander status and child mental health.[6] | Families with socioeconomic disadvantage have higher risk of experiencing racial discrimination.[10] | Children from socioeconomically disadvantaged families are more likely to have mental health and sleep problems than their peers from non-disadvantaged families.[18,19] |

**Supplementary file 3.** Sensitivity analysis including more intermediate confounders in LSAC

To check our results whether are robust or not, we additionally included another five intermediate confounders at 6-7 years in LSAC analysis, given these variables are only available in LSAC.

eTable 2.1 Measures used to define another five intermediate confounders in LSAC

| **Indicator** | **Measurement** | **Example item** | **Coding** |
|---|---|---|---|
| Household member mental illness | The six-item K-6 Depression Scale reported by Parent 1 (P1) and Parent 2 (P2). | "During the past 30 days, about how often did you feel hopeless?" | Score over 13 (mental disorder very likely) categorized as high psychological distress. Neither parent high distress=0; P1 and/or P2 high distress=1. |
| Household member substance abuse | A single-item that asked about parent legal problems, reported by P1. | "In the last year, have any of the following happened to you? Someone in your household had an alcohol or drug problem." | No=0; Yes=1. |
| Family violence | A single item from an adapted version of the Quality of Co-parental Interaction Scale, reported by P1 and P2. | "How often do you have arguments with your partner that end up with people pushing, hitting, kicking or shoving?" | 'Never'=0, 'Rarely' to 'Always'=1. Single parent was coded as 0. |
| Child disability | A single item that asked about the study child's health condition, reported by P1. | "Does the study child have a condition or disability that has lasted for 6 months or more?" | No=0; Yes=1. |
| Residential remoteness | A single item that asked about the region of residence, reported by P1. | "What is your region of residence?" | Metropolitan=0; Not metropolitan=1. |

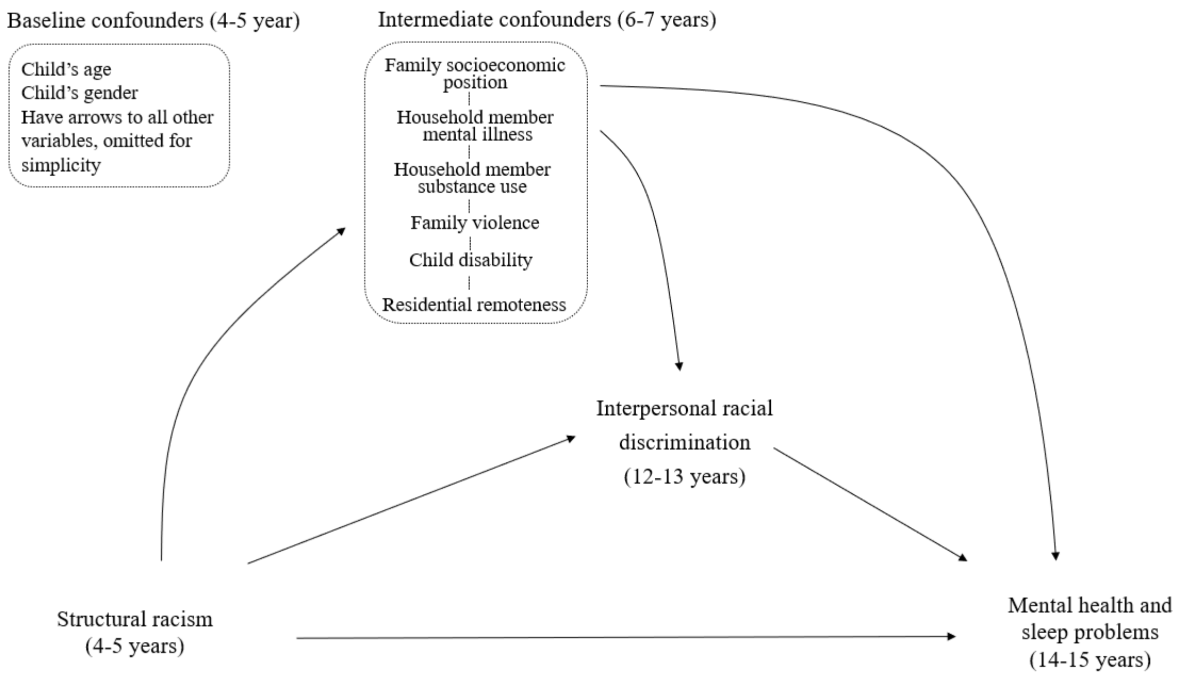


eFigure 2. Conceptual model that depicts the relationship between Aboriginal and Torres Strait

Islander status (as proxies for structural racism exposure; not race and ethnicity as biological or innate constructs) and mental health and sleep problems via interpersonal racial discrimination.

eTable 2.2 Distribution of each intermediate confounder in LSAC. Observed data is shown.

| Variable | LSAC (N=8627) | | |
|---|---|---|---|
| | Frequency (%) / Mean(±SD) | Aboriginal status | |
| | | No | Yes |
| ***Intermediate confounder*** | | | |
| Family socioeconomic position | | | |
| Non-disadvantaged | 5635 (75.0) | 5511 (76.3) | 124 (42.9) |
| Disadvantaged | 1879 (25.0) | 1714 (23.7) | 165 (57.1) |
| Household member mental health illness | | | |
| No | 7193 (96.8) | 6930 (97.0) | 263 (92.9) |
| Yes | 236 (3.2) | 216 (3.0) | 20 (7.1) |
| Household member substance use | | | |
| No | 6490 (97.3) | 6276 (97.5) | 214 (93.9) |
| Yes | 178 (2.7) | 164 (2.5) | 14 (6.1) |
| Family violence | | | |
| No | 7085 (94.0) | 6818 (94.2) | 267 (89.6) |
| Yes | 453 (6.0) | 422 (5.8) | 31 (10.4) |
| Child disability | | | |
| No | 6649 (88.2) | 6402 (88.4) | 247 (82.9) |
| Yes | 891 (11.8) | 840 (11.6) | 51 (17.1) |
| Residential remoteness | | | |
| No | 4308 (57.2) | 4203 (58.1) | 105 (35.2) |
| Yes | 3223 (42.8) | 3030 (41.9) | 193 (64.8) |

LSAC, Longitudinal Study of Australian Children; SD, Standard Deviation.

eTable 2.3 Generalized estimating equations showing the associations between Aboriginal and Torres Strait Islander status, racial discrimination, and mental health and sleep problems, using multiply imputed data for the full cohort of participants (LSAC, N=8627).

| Models | LSAC (RR and its 95% CI) | |
|---|---|---|
| | Unadjusted | Adjusted* |
| ***Association with elevated mental health symptoms*** | | |
| Aboriginal and Torres Strait Islander status (Ref=no) | 3.46 (2.7, 4.44) | 3.45 (2.7, 4.43) |
| Racial discrimination (Ref=no) | 2.74 (2.2, 3.41) | 2.23 (1.77, 2.8) |
| ***Association with sleep problems*** | | |
| Aboriginal and Torres Strait Islander status (Ref=no) | 1.56 (1.05, 2.33) | 1.56 (1.05, 2.32) |
| Racial discrimination (Ref=no) | 1.64 (1.28, 2.1) | 1.56 (1.2, 2.02) |
| ***Association between racial discrimination*** | | |
| Aboriginal status (Ref=no) | 3.36 (2.55, 4.44) | 3.35 (2.54, 4.42) |

CI, confidence interval; LSAC, Longitudinal Study of Australian Children; Ref, reference group; RR, risk ratio.
* Adjusted confounders were child's age and gender. Models for the association between racial discrimination and each outcome were additionally adjusted for Aboriginal and Torres Strait Islander status, family socioeconomic position, household member mental illness, household member substance abuse, family violence, child disability, residential remoteness.

eTable 2.4 Results from causal mediation analysis: Estimated effects on the prevalence of mental health and sleep problems by hypothetical interventions eliminating racial discrimination in Aboriginal and Torres Strait Islander children, using multiply imputed data for the full cohorts in LSAC.

| Effect | LSAC (N=8627) | | |
|---|---|---|---|
| | Estimate of absolute risk reduction (%) 95% CI | Proportion of inequities eliminated | Remaining inequities (%) 95% CI |
| ***Mental health problems*** | | | |
| Total adjusted marginal prevalence difference | 23.2 (18.8, 27.6) | - | - |
| Risk reduction from intervening on racial discrimination | 3.3 (0.1, 6.5) | 14.2 | 19.9 (14.7, 25.1) |
| ***Sleep problems*** | | | |
| Total adjusted marginal prevalence difference | 6.2 (0, 12.5) | - | - |
| Risk reduction from intervening on racial discrimination | -0.5 (-4.4, 3.5) | 0 | 6.7 (-0.8, 14.2) |

CI, confidence interval. Baseline confounders controlled for were child's age and gender. Intermediate confounder controlled for was family socioeconomic position, household member mental health, household member substance use, family violence, child disability and remoteness.

**Supplementary file 4:** Additional details about the outcome and mediator models used for estimating the interventional effects of hypothetical interventions using extended G-computation.

In the primary analysis (adjusted for age, sex and intermediate confounder socioeconomic position) the outcome model included the exposure (Aboriginal and Torres Strait Islander status), mediator (racial discrimination), intermediate confounder (socioeconomic position), baseline confounders (age and sex), and two-way interactions between exposure and baseline confounders, exposure and mediator, exposure and intermediate confounder and, mediator and intermediate confounder. The model for the mediator included exposure, baseline confounders, and two-way interactions between exposure and baseline confounders, while the model for intermediate confounder included the exposure, baseline confounders, and two-way interactions between the exposure and baseline confounders.

In the analysis in LSAC including additional intermediate confounders, the outcome model included the exposure, mediator, baseline confounders, intermediate confounders (socioeconomic position, household member mental health, household member substance use, family violence, child disability and remoteness) and the interactions between exposure and baseline confounders, exposure and mediator, exposure and intermediate confounders, mediator and intermediate confounders. As in the primary analysis, the mediator model included exposure, baseline confounders, and two-way interactions between exposure and baseline confounders. The joint distribution of the intermediate confounders was modelled by decomposing the joint distribution into sequential conditional distributions assuming a non-causal order as follows:

- Regression for $L_6$ on exposure, baseline confounders and two-way interactions between exposure and baseline confounders
- Regression for $L_5$ on exposure, baseline confounders, $L_6$ and two-way interactions between exposure and baseline confounders, and exposure and $L_6$
- Regression for $L_4$ on exposure, baseline confounders, $L_6$, $L_5$, and two-way interactions between exposure and baseline confounders, and exposure and $L_6$, and exposure and $L_5$
- Regression for $L_3$ on exposure, baseline confounders, $L_6$, $L_5, L_4$, and two-way interactions between exposure and baseline confounders, and exposure and $L_6$, and exposure and $L_5$, and exposure and $L_4$
- Regression for $L_2$ on exposure, baseline confounders, $L_6$, $L_5, L_4, L_3$, and two-way interactions between exposure and baseline confounders, and exposure and $L_6$, and exposure and $L_5$, and exposure and $L_4$, and exposure and $L_3$
- Regression for $L_1$ on exposure, baseline confounders, $L_6$, $L_5, L_4, L_3, L_2$, and two-way interactions between exposure and baseline confounders, and exposure and $L_6$, and exposure and $L_5$, and exposure and $L_4$, and exposure and $L_3$, and exposure and $L_2$

Where $L_1$: socioeconomic position, $L_2$: household member mental health, $L_3$: household member substance use, $L_4$ :family violence, $L_5$ : child disability and , $L_6$: remoteness

### References


1. Priest N, Chong S, Truong M, et al. *Findings from the 2017 Speak Out Against Racism (SOAR) student and staff surveys. CSRM working paper no. 3/2019*. 2019. https://csrm.cass.anu.edu.au/sites/default/files/docs/2019/8/CSRM-WP-SOAR_PUBLISH_1.pdf
2. Shahaeian A, Wang C, Tucker-Drob E, Geiger V, Bus AG, Harrison LJ. Early Shared Reading, Socioeconomic Status, and Children's Cognitive and School Competencies: Six Years of Longitudinal Evidence. *Scientific Studies of Reading*. 2018:1-18.
3. Priest N, King T, Bécares L, Kavanagh A, M. Bullying victimization and racial discrimination among Australian children. Article. *American Journal of Public Health*. 2016;106(10):1882-1884. doi:10.2105/AJPH.2016.303328
4. Australian Institute of Health and Welfare. *Australia's welfare 2017: in brief. Cat. no. AUS 215*. 2017.
5. De Maio JA, Zubrick SR, Silburn SR, et al. The Western Australian Aboriginal child health survey: Measuring the social and emotional wellbeing of Aboriginal children and intergenerational effects of forced separation. *Perth: Curtin University of Technology and Telethon Institute for Child Health Research*. 2005;
6. Williamson A, Gibberd A, Hanly MJ, et al. Social and emotional developmental vulnerability at age five in Aboriginal and non-Aboriginal children in New South Wales: a population data linkage study. *International Journal for Equity in Health*. 2019/07/31 2019;18(1):120. doi:10.1186/s12939-019-1019-x
7. Blunden S, Fatima Y, Yiallourou S. Sleep health in Indigenous Australian children: a systematic review. *Sleep medicine*. 2021;80:305-314.
8. Benner AD, Graham S. The antecedents and consequences of racial/ethnic discrimination during adolescence: does the source of discrimination matter? *Developmental Psychology*. Aug 2013;49(8):1602-13. doi:10.1037/a0030557
9. Priest N, Chong S, Truong M, et al. Racial discrimination and socioemotional and sleep problems in a cross-sectional survey of Australian school students. *Archives of Disease in Childhood*. Jul 28 2020;105(11):1079-1085. doi:https://doi.org/10.1136/archdischild-2020-318875
10. Shepherd CCJ, Li J, Cooper MN, Hopkins KD, Farrant BM. The impact of racial discrimination on the health of Australian Indigenous children aged 5–10 years: analysis of national longitudinal data. journal article. *International Journal for Equity in Health*. July 03 2017;16(1):116. doi:10.1186/s12939-017-0612-0
11. Thurber K, Walker J, Dunbar T, et al. *Measuring child mental health, psychological distress, and social and emotional wellbeing in the longitudinal study of indigenous children*. 2019.
12. Australian Mental Health Outcomes and Classification Network. *Strengths and difficulties questionnaire: Training manual*. 2005. https://www.amhocn.org/sites/default/files/publication_files/sdq_manual_0.pdf
13. Kelly Y, Zilanawala A, Booker C, Sacker A. Social media Use and adolescent mental health: findings from the UK Millennium Cohort Study. *EClinicalMedicine*. 2018/12/01/ 2018;6:59-68. doi:10.1016/j.eclinm.2018.12.005
14. Evans-Whipp T, Gasser C. *Are children and adolescents getting enough sleep?*

2018:29-46. *Growing Up in Australia: The Longitudinal Study of Australian Children (LSAC), Annual Statistical Report*.
15. Lawrence D, Johnson S, Hafekost J, et al. The mental health of children and adolescents: Report on the second Australian Child and Adolescent Survey of Mental Health and Wellbeing. 2015;
16. Chen Y-L, Tseng W-L, Yang L-K, Gau SS-F. Gender and Age Differences in Sleep Problems in Children: Person-Oriented Approach With Multigroup Analysis. *Behavioral Sleep Medicine*. 2019/05/04 2019;17(3):302-313. doi:10.1080/15402002.2017.1357117
17. Blakemore T, Strazdins L, Gibbings J. Measuring family socioeconomic position. *Australian Social Policy*. 2009;8:121-168.
18. Reiss F. Socioeconomic inequalities and mental health problems in children and adolescents: A systematic review. *Social Science & Medicine*. 2013;90(0):24-31. doi:http://dx.doi.org/10.1016/j.socscimed.2013.04.026
19. Park JW, Hamoda MM, Almeida FR, et al. Socioeconomic inequalities in pediatric obstructive sleep apnea. *Journal of Clinical Sleep Medicine*. 2021:jcsm-9494.
